# Policy Levers of HIV Control: Targeted Service Coverage, Financial Protection, and Estimated New HIV Infections in Southeast Asia, 2013–2022

**DOI:** 10.64898/2026.04.11.26350590

**Authors:** Jason Hung, Adrian Smith

## Abstract

The global ambition to end the human immunodeficiency virus (HIV) epidemic requires understanding which system-level policy levers, enacted under the framework of Universal Health Coverage (UHC), are most effective in achieving both transmission reduction and increasing access to diagnosis and treatment. This study addresses an important evidence gap by quantifying the within-country association between measurable UHC policy indicators and the estimated rate of new HIV infections across nine Southeast Asian countries between 2013 and 2022. Employing a Fixed-Effects panel data methodology, the analysis controls for time-invariant national heterogeneity, so that the estimates describe change over time within each country. We found that within-country changes over time in total current health expenditure (CHE) as a percentage of gross domestic product (GDP) were not statistically significantly associated with the estimated rate of new HIV infections. However, increases in the UHC Infectious Disease Service Coverage Index were statistically significantly associated with concurrent reductions in HIV incidence (*p* < 0.001), suggesting that targeted, disease-specific service delivery is the most consistent of the policy levers we examined. In addition, the UHC Reproductive, Maternal, Newborn, and Child Health Service Coverage Index exhibited a statistically significant positive association with HIV incidence (*p* < 0.01). We cannot determine whether this positive association indicates improved detection of previously undiagnosed cases, a feature of how UNAIDS estimates incidence, or the influence of the wider sexual and reproductive health system. Furthermore, out-of-pocket (OOP) health expenditure as a percentage of CHE showed a counterintuitive negative association with the estimated rate of new HIV infections (*p* < 0.01). Because this metric measures spending across the whole health system, further research at the individual level is needed before this finding can inform policy. These findings suggest that domestic HIV policymakers should prioritise targeted investment in infectious disease services over aggregate fiscal commitment.

## Introduction

National control of the human immunodeficiency virus (HIV) epidemic has long been a leading global public health imperative (Kumah et al., 2023). Despite significant advances in biomedical prevention and treatment, notably Pre-Exposure Prophylaxis (PrEP), and the success of Treatment as Prevention, the epidemic persists, and in the absence of cure requires continuous, targeted interventions integrated within resilient national health systems (Beyrer et al., 2025). Globally, public health efforts now focus on achieving and sustaining ambitious treatment goals—the UNAIDS 95-95-95 targets, aiming for 95% of people living with HIV to know their status, 95% of those diagnosed receiving antiretroviral therapy (ART), and 95% of those on ART achieving viral suppression (Frescura et al., 2022).

Southeast Asia presents a heterogeneous public health landscape in this regard, with certain countries, such as Thailand, being recognised as regional exemplars for their progress towards these goals. By the end of 2022, 90% of the Thai population living with HIV was aware of their status, 90% of those diagnosed were on ART, and 97% of those on treatment achieved a suppressed viral load (UNAIDS, 2024). However, many countries in the region face ongoing challenges related to access, adherence, and financial protection, underpinning the necessity for robust research into effective system-level policy measures. As of the end of 2021, neighbouring countries such as Indonesia (28%), the Philippines (41%), Malaysia (55%), and Laos (57%) reported relatively low ART treatment coverage for HIV (UNAIDS Asia-Pacific, 2023) (specifically referring to the estimated percentage of people living with HIV receiving ART).

Tackling these entrenched disparities and ensuring universal access to ART requires leveraging comprehensive, system-level reform. In this context, Universal Health Coverage (UHC) serves as the internationally mandated vehicle for strengthening health systems and achieving long-term infectious disease control (i.e., curbing new HIV infections) and population health (Kieny et al. 2017). Endorsed by the *2018 Astana Declaration*, UHC mandates integrated health services, with primary health care forming the foundational core (WHO, 2018). The integration of disease-specific programmes into wider health system strengthening under UHC has been uneven across regions. Ooms and Kruja (2019) argue that integrating the HIV response into UHC is difficult in practice, and in many settings HIV programmes have remained organised separately from the wider health system and dependent on external financing. For HIV, this shift recognises that while the availability of ART is important, sustained success requires addressing system-level barriers. Research indicates that long wait times, inadequate patient follow-up, and logistical hurdles related to fragmented health systems discourage adherence and patient retention (Alrasheedi et al., 2019; Kagee et al., 2011; Kogi et al., 2024). Suboptimal coverage of universal ART programmes risks contributing to avoidable transmission and drug resistance (WHO, 2025), highlighting the need for efficient, well-capacitated health systems inherent to the UHC framework.

The primary policy debate surrounding UHC implementation focuses on the empirical efficacy of two core economic levers, namely fiscal commitment and patient financial protection. Regarding resource allocation, the literature suggests that increased governmental priority (measured by total health expenditure relative to GDP) should translate into greater resources for prevention and treatment (Bein & Coker-Farrell, 2020), theoretically leading to reduced HIV incidence. This is contrasted by the scholarly debate on patient financial protection, where the global health literature conventionally argues that high OOP payments pose significant financial barriers, often leading to forgone treatment, diminished compliance, and poorer outcomes (Koskinen et al., 2019; Xu et al., 2003). Crucially, out-of-pocket (OOP) expenditure covers spending across the whole health system, not spending on HIV services alone, and existing macroeconomic studies do not separate the cost of care for people already diagnosed from the deterrent effect of those costs on people not yet tested.

This empirical ambiguity extends to the dynamic implementation of service coverage. While the UHC Infectious Disease Service Coverage Index quantifies the effectiveness of services essential for managing diseases like HIV, providing a clear expectation of transmission reduction, a significant gap remains in understanding what the reproductive and maternal health index measures in relation to HIV. The UHC Reproductive, Maternal, Newborn, and Child Health Service Coverage Index, though theoretically linked to transmission reduction through sexual health and prevention of mother to child transmission initiatives (An et al., 2015), carries the potential to function as an indicative surveillance metric. It is therefore important to establish whether service expansion in Reproductive, Maternal, Newborn, and Child Health primarily drives actual HIV transmission reduction or, in a counter-intuitive finding, enhances case identification and reporting. We accordingly approached the analysis with three prior expectations, namely that greater fiscal commitment and greater infectious disease service coverage would each be associated with lower estimated HIV incidence, that a greater OOP burden would be associated with higher estimated HIV incidence, and that the direction of any association with reproductive, maternal, newborn, and child health coverage could not be predicted in advance, given that a single index may capture transmission reduction and enhanced case identification simultaneously. This paper focuses on quantifying the association between several measurable policy indicators—including health financing commitment (current health expenditure (CHE) as a percentage of gross domestic product (GDP)), financial burden on individuals (OOP expenditure as a percentage of CHE), and the effective implementation of coverage across various essential services—and the rate at which new HIV infections occur.

Analyses of policy indicators in this region have generally not accounted for intrinsic heterogeneity between countries, and the within-country association between specific health policy levers and estimated HIV incidence remains underexplored across Southeast Asian economies. This longitudinal panel analysis addresses this gap by employing a robust Fixed-Effects panel data methodology spanning nine Southeast Asian countries over the period 2013 to 2022 (Appendix 1). By quantifying the within-country impact of each policy lever, the analysis establishes the empirical context for future micro-level investigations. Our research aim is to determine which time-variant health policy factors, derived from World Health Organisation (WHO) data, demonstrate a statistically significant association with the estimated rate of new

HIV infections in the general population, ensuring that the analysis controls for time-invariant national differences.

## Materials and Methods

### Data Extraction and Variable Selection

In this study, we extracted data for all variables included in this panel analysis using Python to interface with the WHO’s Global Health Observatory (GHO) Application Programming Interface (API) ensuring access to the most current WHO-verified data. The API-based approach enabled systematic extraction of time-series country level data covering the period from 2013 to 2022 for all Southeast Asian countries.

Table 1 details all variables used for panel data analysis. The primary outcome of interest is the estimated number of new HIV infections per 1,000 uninfected population per year in each country, *hiv_incidence*_*i*_,_*t*_ (Table 1). These estimates are produced using the Joint United Nations Programme on HIV/AIDS (UNAIDS)-supported Spectrum mathematical modelling software package. The software package was developed by the Futures Institute; and the HIV estimates are reviewed and validated in partnership with WHO and United Nations Children’s Fund (UNAIDS, 2014). The denominator for *hiv_incidence*_*i*_,_*t*_ is the population not living with HIV in each country and year, which UNAIDS derives within Spectrum from United Nations population estimates.

**Table 1:** Description of Variable Definitions.

| Variable | Description |
| --- | --- |
| $country_i$ | The code represents the country. $i$ is the country index (**Group Identifier**) |
| $year_t$ | The calendar year of the data point. $t$ is the time index (**Time Identifier**) |
| $hiv\_incidence_{i,t}$ | Estimated number of new HIV infections per 1,000 uninfected population per year (**Dependent Variable**) |
| $health\_infrastructure\_index_i$ | A time-invariant composite measure (hence, no $t$ subscript) of the country's overall healthcare system capacity, which includes metrics measuring the density of healthcare facilities and personnel (**Time-Invariant Explanatory Variable**) |
| $che\_gdp_{i,t}$ | Total CHE from all financing sources as a percentage of GDP, compiled by WHO under the System of Health Accounts 2011 framework. Modelled as a share and not an amount, so that fiscal commitment is expressed relative to national economic capacity (**Time-Variant Explanatory Variable**) |
| $oop\_che_{i,t}$ | OOP health expenditure as a percentage of the CHE, measuring the share of all health spending that households pay directly at the point of care. This covers the whole health system and is not specific to HIV services (**Time-Variant Explanatory Variable**) |
| $uhc\_infect\_sci_{i,t}$ | A WHO subindex of the SDG 3.8.1 UHC Service Coverage Index, calculated as a population-weighted geometric mean of four coverage measures, namely tuberculosis case detection and treatment, ART coverage, insecticide-treated net use in malaria-endemic areas, and use of at least basic sanitation. The index ranges from 0 to 100, where 100 is full coverage (**Time-Variant Explanatory Variable**) |
| $uhc\_rmnch\_sci_{i,t}$ | A WHO subindex of the SDG 3.8.1 UHC Service Coverage Index, calculated in the same way from four coverage measures, namely demand for family planning satisfied by modern methods, four or more antenatal care visits, three doses of diphtheria-tetanus-pertussis containing vaccine, and care-seeking for suspected childhood acute respiratory infection. None of the four records HIV testing. The index ranges from 0 to 100 (**Time-Variant Explanatory Variable**) |

Source: WHO's GHO

The explanatory variables used to model this outcome are detailed in Table 1. Because each of these variables is a ratio or a composite that WHO constructs from other measures, we set out below what each one measures before introducing its notation. Total CHE as a percentage of GDP, denoted *che_gdp*_*i*_,_*t*_, is compiled by WHO within the Global Health Expenditure Database under the System of Health Accounts 2011 framework. This metric measures the share of national economic output devoted to health from all financing sources, whether governmental, private, or external.

OOP health expenditure as a percentage of CHE, denoted *oop_che*_*i*_,_*t*_, is compiled from the same accounting framework and measures the proportion of all health spending that households pay directly at the point of care. This metric covers spending across the whole health system and is not specific to HIV services, since households spend on every category of health care and HIV services form only a small part of that spending in Southeast Asia. Any association we estimate between this metric and HIV incidence is therefore an association between national aggregates and not evidence about the costs individual patients face when seeking HIV care.

The UHC Infectious Disease Service Coverage Index, denoted *uhc_infect_sci*_*i*_,_*t*_, is one of four subindices from which WHO constructs the SDG 3.8.1 UHC Service Coverage Index. WHO calculates this subindex as a population-weighted geometric mean of four underlying coverage measures, each scaled from 0 to 100, namely the percentage of incident tuberculosis cases detected and treated, the percentage of adults and children living with HIV receiving ART, the percentage of the population in malaria-endemic areas sleeping under an insecticide-treated net on the previous night, and the percentage of the population using at least basic sanitation services. This subindex therefore measures the delivery of targeted, disease-specific services, and it is not a measure of how far health services are integrated with one another.

The UHC Reproductive, Maternal, Newborn, and Child Health Service Coverage Index, denoted *uhc_rmnch_sci*_*i*_,_*t*_, is constructed in the same way from four coverage measures, namely the percentage of women of reproductive age whose demand for family planning is satisfied by modern methods, the percentage of women with a live birth who received antenatal care on four or more occasions, the percentage of infants receiving three doses of diphtheria-tetanus-pertussis containing vaccine, and the percentage of children under five with symptoms of acute respiratory infection for whom advice or treatment was sought. None of these four measures records HIV testing, HIV diagnosis, or the prevention of mother-to-child transmission, so this index serves only as a proxy for the reach of the reproductive and maternal health services through which HIV testing is commonly delivered.

Finally, geographical coverage was restricted to nine Southeast Asian countries (Appendix 1), as Brunei was ultimately excluded from the analysis. The GHO API consistently reported no available data points for *hiv_incidence*_*i*_,_*t*_ for Brunei across the entire 2013-2022 period. Including Brunei would have resulted in its complete exclusion through listwise deletion in every regression, thus complicating the panel structure unnecessarily. Its exclusion ensures a cleaner, more reliable dataset for the remaining nine countries.

The primary measures of healthcare capacity employed in this study are *health_posts_density*_*i*_,_*t*_ (access to primary care) and *hospitals_density*_*i*_,_*t*_ (advanced capacity), both sourced from the GHO. Given a high degree of positive collinearity between these two metrics, a single composite *health_infrastructure_index*_*i*_ was constructed to avoid multicollinearity. We performed *Z*-score standardisation on both variables to ensure equal contribution despite their disparate scales. The final *health_infrastructure_index*_*i*_ score was calculated as the average of the two standardised *Z*-scores, or, if only one component was available, that single standardised score was used for the index value. This constructed index shows a fixed, baseline structural capacity of the country’s health system.

### Data Imputation

The GHO does not report every variable annually for every country. Gaps occurred in *health_posts_density*_*i*_,_*t*_ and *hospitals_density*_*i,t*_, and they follow how often WHO reports these measures, and are not country-specific non-response. To maintain statistical power and to avoid the selection bias that listwise deletion would introduce in a panel of only nine countries, we carried the last recorded value forward until a new value was observed, following the Last Observation Carried Forward convention described by Little and Rubin (2019). We applied this convention only to the two facility density measures, which long-term planning and construction cycles determine and which therefore move slowly from one year to the next, so that the value from the preceding year is a reasonable proxy for the missing period. We acknowledge, following Lachin (2016), that carrying values forward reduces the year-to-year variation within a country, so that where the density of hospitals and health posts actually changed in the years without a reported value, our standard errors will be misleading.

### Modelling

When building linear regression models for panel data, we used *hiv_incidence*_*i*_,_*t*_ as the dependent variable and *health_infrastructure_index*_*i*_, *che_gdp*_*i*_,_*t*_, *oop_che*_*i*_,_*t*_, *uhc_infect_sci*_*i*_,_*t*_, and *uhc_rmnch_sci*_*i*_,_*t*_ as the independent variables (in Table 3). Here are the regression equations and assumptions for the Fixed-Effects and Random-Effects models:

#### (1) Fixed-Effects Model

The Fixed-Effects model (or “within” estimator) controls for all unobserved differences between countries by removing the country-specific mean, so as to isolate the effect of changes within each country over time.

*Fixed-Effects Model: hiv_incidence*_*i,t*_ *= β*_*0*_ *+ β*_*2*_*che_gdp*_*i,t*_ *+ β*_*3*_*oop_che*_*i,t*_ *+ β*_*4*_ *uhc_infect_sci*_*i,t*_ *+ β*_*5*_*uhc_rmnch_sci*_*i,t*_ *+ α*_*i*_ *+ ϵ*_*it*_

Assumptions for the Fixed-Effects estimation:

1. **Strict exogeneity**: The time-varying error term ϵ_it_ is uncorrelated with the regressors in all time periods: *E[ϵ*_*it*_ | *X*_*i1*_, …, _*XiT*_, *α*_*i*_] = 0.
2. **No perfect collinearity**: The time-varying regressors (X_it_) must vary over time within each country.
3. **Time-invariant variable omission**: The time-invariant variable (Z_i_) is absorbed into the unobserved country-specific effect (α_i_) and cannot be estimated (hence it is omitted in the Fixed-Effects column).
4. **Error structure**: The idiosyncratic errors (ϵ_it_) are assumed to be independent across observations, but because *vce(robust)* was used, the model accommodates heteroscedasticity (non-constant variance) in the errors.

#### (2) Random-Effects Model

The Random-Effects model assumes that the unobserved country-specific effects (α_i_) are random and uncorrelated with the predictors. This model allows for the estimation of time-invariant variables (Z_i_).

*Random-Effects Model: hiv_incidence*_*i,t*_ *= β*_*0*_ *+ β*_*1*_*health_infrastructure_index*_*i*_ *+ β*_*2*_*che_gdp*_*i,t*_ *+ β*_*3*_*oop_che*_*i,t*_ *+ β*_*4*_ *uhc_infect_sci*_*i,t*_ *+ β*_*5*_*uhc_rmnch_sci*_*i,t*_ *+ (α*_*i*_ *+ ϵ*_*it*_)

Assumptions for the Random-Effects estimation:

1. **No correlation:** The country-specific error term (α_i_) must be uncorrelated with all explanatory variables (X_it_ and Z_i_). This assumption is tested by the Sargan-Hansen C-statistic.
2. **Error structure:** The composite error term (u_it_ = α_i_ + ϵ_it_) is assumed to be independent across countries.
3. **Heteroscedasticity**: As *vce(robust)* was used, the estimation accommodates Heteroscedasticity in the errors (ϵ_it_ and α_i_).

We also re-ran the Fixed-Effects and Random-Effects models with the explanatory variables lagged by one year and with standard errors clustered at the country level, to check whether serial autocorrelation (i.e., where observations within a country are correlated across successive time periods) had produced spurious significance in the models reported in Table 3. The specifications, assumptions and results for these lagged models are reported in the Supplementary Material (Supplementary Table 1). To choose between the Fixed-Effects and Random-Effects estimators, we conducted a Sargan-Hansen test.

We assessed whether there was any collinearity issue among the independent variables. Appendix 2 shows the variance inflation factor (VIF) values of all independent variables that are utilised for building linear regression models for panel data. The VIF values in the panel dataset indicate that there is no significant multicollinearity among the independent variables, as values fell below the conventional threshold of 5.0, indicating acceptable collinearity (Hair et al., 2010). Table 2 shows the correlations between all independent variables that are utilised for regression analysis (in Table 3 and Supplementary Table 1). Several variable pairs exhibited moderate to strong bivariate correlations at a statistical significance (*p* < 0.05) level (Table 2). While the correlations between *oop_che*_*i,t*_ and *health_infrastructure_index*_*i*_ (*ρ* = −0.56; *p* < 0.001) and between *uhc_rmnch_scii*_,*t*_ and *uhc_infect_scii*_,*t*_ (*ρ* = 0.66; *p* < 0.001) are both fairly strong, the VIF test results (in Appendix 2) already assure that no multicollinearity issues need to be addressed.

**Table 2:** Spearman Rank Correlation Table between Independent Variables.

|  | *health_infrastructure_index<sub>i</sub>* | *che_gdp<sub>i,t</sub>* | *oop_che<sub>i,t</sub>* | *uhc_infect_sci<sub>i,t</sub>* | *uhc_rmnh_sci<sub>i,t</sub>* |
| --- | --- | --- | --- | --- | --- |
| *health_infrastructure_index<sub>i</sub>* | 1.0000 |  |  |  |  |
| *che_gdp<sub>i,t</sub>* | -0.1259 | 1.0000 |  |  |  |
| *oop_che<sub>i,t</sub>* | -0.5571*** | 0.2374* | 1.0000 |  |  |
| *uhc_infect_sci<sub>i,t</sub>* | 0.2554* | 0.3685*** | -0.4754*** | 1.0000 |  |
| *uhc_rmnh_sci<sub>i,t</sub>* | 0.3363** | 0.1116 | -0.6615*** | 0.6567*** | 1.0000 |

N (Observations) = 80
Note: \*\*\*, \*\*, \* denotes significance at the 0.1%, 1%, and 5% level, respectively.

**Table 3:** Regression Results for HIV Incidence (Fixed-Effects vs. Random-Effects with Robust Specification Test)

| Variable | Fixed-Effects model |  | Random-Effects model |  |
| --- | --- | --- | --- | --- |
|  | coefficient | standard error | coefficient | standard error |
| *health_infrastructure_index<sub>i</sub>* | / |  | -0.0250 | 0.0418 |
| *che_gdp<sub>i,t</sub>* | -0.0024 | 0.0082 | 0.0001 | 0.0060 |
| *oop_che<sub>i,t</sub>* | -0.0036** | 0.0016 | -0.0012 | 0.0009 |
| *uhc_infect_sci<sub>i,t</sub>* | -0.0028*** | 0.0008 | -0.0019** | 0.0008 |
| *uhc_rmnch_sci<sub>i,t</sub>* | 0.0074** | 0.0023 | 0.0020 | 0.0017 |
| constant | -0.0744 | 0.1471 | 0.1726* | 0.0991 |
| N (Observations) | 90 |  | 80 |  |
| N (Groups) | 9 |  | 8 |  |
| F-statistic/ Wald $\chi^2$ | F(4,8) = 6.67 | | $\chi^2(5) = 8.14$ | |
| p-value | 0.0116 |  | 0.1486 |  |
| Sargan-Hansen C-statistic | $\chi^2(4) = 48.81$ | $p < 0.001$ | | |

Note: \*\*\*, \*\*, \* denotes significance at the 0.1%, 1%, and 5% level, respectively.
Note: The lower N in the Random-Effects model reflects the exclusion of Vietnam, for which the *health\_infrastructure\_index* could not be estimated due to missing facility density data throughout the study period.

To select between the Fixed-Effects and Random-Effects estimators, we employed the Sargan-Hansen C-statistic (Arellano, 1993; Schaffer & Stillman, 2006), which, unlike the conventional Hausman test, remains valid when robust or clustered standard errors are used to accommodate the heteroscedasticity and within-country autocorrelation found in national health data (Wooldridge, 2010). This Sargan-Hansen C-statistic test assesses the validity of the central assumption of the Random-Effects model, namely that the unobserved country-specific effects are uncorrelated with the regressors.

## Results

Across the nine Southeast Asian countries over the 2013–2022 period, the overall regional incidence rate declined from 0.19 per 1,000 uninfected population per year in 2013 to 0.13 in 2022, however there was between-country heterogeneity. A test for trend across the 10 years did not reach statistical significance (Fixed-Effects linear time trend, *β* = −0.0045 per year, 95% CI = −0.0095 to 0.0005, *p* = 0.07). Most countries, including Thailand, Cambodia, and Singapore, showed lower HIV incidence at the end of the decade than at the beginning. Conversely, the Philippines had an upward trend in new infections across the study period, highlighting the diverse epidemiological stages present in the region. The high degree of both cross-sectional and temporal variation in the outcome variable suggests that the dataset is suitable for measuring the within-country impact of time-variant policy changes, which is the aim of the subsequent regression analyses.

Table 3 shows the empirical outputs of the Fixed-Effects model, Random-Effects model, and Sargan-Hansen C-statistic. The application of the Sargan-Hansen C-statistic to the final Random-Effects estimation yielded a test statistic of 48.81 with four degrees of freedom (*p* < 0.001), suggesting that the Fixed-Effects assumption is held, prompting its selection as the preferred and consistent estimator for interpretation.

The coefficients in Table 3 describe change over time within a country, because the Fixed-Effects estimator removes each country’s mean and uses only the within-country variation across the 10 years of the panel. They are not comparisons between one country and another. After adjustment for the other covariates, a one percentage point increase in OOP health expenditure as a percentage of CHE was associated with a decrease of 0.0036 in the estimated number of new HIV infections per 1,000 uninfected population per year (SE = 0.0016, *p* < 0.01). A one-point increase on the UHC Infectious Disease Service Coverage Index was associated with a decrease of 0.0028 per 1,000 uninfected population per year (SE = 0.0008, *p* < 0.001). A one-point increase on the UHC Reproductive, Maternal, Newborn, and Child Health Service Coverage Index was associated with an increase of 0.0074 per 1,000 uninfected population per year (SE = 0.0023, *p* < 0.01).

When the policy variables were lagged by one year, no empirical findings retained statistical significance at the 0.05 or 0.10 level (Supplementary Table 1). We report this result without treating the loss of significance as evidence that the associations in Table 3 were contemporaneous, because a panel of nine countries has limited statistical power to detect an association between a policy variable in one year and HIV incidence in the following year.

## Discussion

The Fixed-Effects design allows an indicative evaluation of policy levers irrespective of fixed national characteristics such as underlying health culture (Airhihenbuwa & Webster, 2004), political stability (Menon-Johansson, 2005), or baseline structural capacity (Stannah et al., 2024). Within this framework, the analysis suggests that increases in the UHC Infectious Disease Service Coverage Index were significantly associated with concurrent reductions in HIV incidence. Such evidence offers empirical support for the effectiveness of targeted, disease-specific service delivery but not for service integration as such, given that this subindex is constructed from measures of coverage for individual disease programmes. This aligns with ecological evidence that high ART coverage is associated with a reduced risk of HIV acquisition at the population level (Tanser et al., 2013). The index does not measure screening, retention in care, or viral suppression directly, so we cannot tell which of them explains the association. Among the levers we examined, targeted service coverage showed a stronger association with HIV incidence than aggregate fiscal commitment, which showed none. We do not interpret the absence of significant associations in the lagged specification as evidence that the effects reported in Table 3 were contemporaneous. Because neither national service coverage nor modelled HIV incidence changes appreciably from one year to the next, we are unable to establish whether the loss of statistical significance under a one-year lag suggests the limited statistical power available from a panel of nine countries, or indicates that the associations reported in Table 3 were themselves spurious.

The negative association between OOP expenditure and HIV incidence is contrary to existing global health literature, which states that high private health spending increases barriers to care (Voto et al., 2025), diminishes compliance (Barennes et al., 2015), and ultimately leads to higher rates of new infection (Sullivan et al., 2025) due to reduced treatment adherence. The OOP metric measures spending across the whole health system, not spending on HIV services alone, and our data cannot establish which explanation accounts for this negative association. Confounding by the wider health system and by related factors such as national income is one possible explanation, since the countries with a higher household share of health spending were also the countries with better-resourced public prevention programmes and with less dependence on external HIV financing. Because the OOP data are national aggregates, we also cannot tell whether higher costs stopped vulnerable, undiagnosed people from seeking HIV test, so that the negative coefficient shows that fewer of those people were diagnosed, and not that fewer were infected. Further research at the individual level is needed before the OOP finding can inform policy.

In addition to the financial associations, our study identifies a significant positive coefficient for the UHC Reproductive, Maternal, Newborn, and Child Health Index. This observation could imply that higher coverage in these services likely enhanced HIV detection, often through routine testing incorporated into antenatal care or sexual health clinics, therefore raising the measured incidence rate without a corresponding rise in HIV transmission. Our data, however, cannot validate such an implication. *hiv_incidence* is an estimate produced by the Spectrum model, not a count of the cases each country diagnosed. Antenatal testing is also provided primarily to prevent mother-to-child transmission, which should reduce incidence. Thus, this study reports the positive coefficient between UHC Reproductive, Maternal, Newborn, and Child Health Index and HIV incidence without determining whether the association indicates enhanced detection, an artefact of incidence estimation, or confounding by the wider sexual and reproductive health system.

These findings support continued national investment in infectious disease service coverage. The targeted infectious disease service coverage metric showed the most consistent association with lower HIV incidence within countries. Among the WHO indicators available, the targeted infectious disease service coverage metric is therefore the most useful for national and regional HIV planning. The reproductive, maternal, newborn, and child health index, by contrast, cannot be read as an indicator of HIV control for national and regional planning.

## Conclusions

Across nine Southeast Asian countries from 2013 to 2022, targeted infectious disease service coverage was the only policy lever consistently associated with lower HIV incidence, while aggregate health spending as a share of GDP was not. Higher reproductive, maternal, newborn, and child health coverage was associated with higher reported incidence, and higher OOP expenditure with lower incidence, but our data cannot establish what either association means. Operational service delivery, and not the overall level of health spending, is where the evidence points for HIV epidemic control in the region. Future research should test these associations with individual-level data and with direct measures of PrEP uptake and viral suppression.

## Limitations and Future Research Directions

The analysis is constrained by several limitations inherent to the structure and availability of global health data. First, the methodological decision of using the Fixed-Effects estimator prevents the quantification of the direct impact of the time-invariant fixed structural health capacity. Second, the study’s reliance on aggregated UHC indices limits the ability to directly measure the core biomedical mechanisms now driving incidence reduction, such as the scaleup of PrEP use or the system’s success in achieving viral suppression (the efficacy of U=U). These factors are highly correlated with the rate of new HIV infections, but suitable longterm national panel data for them are not available via the WHO’s GHO. The UHC Infectious Disease Service Coverage Index therefore serves as a proxy for these biomedical cascades. Third, the decision to rely solely on the rate of new HIV infections is attributed to the severe sparsity and time-invariance of the total percentage of the population living with HIV data (specifically HIV prevalence), limiting the analysis to the rate of new infections only. Fourth, the short time-series nature of the panel dataset (T=10; 2013-22) limits the application of advanced time-series econometrics, such as robust unit root testing. Fifth, because we measured the explanatory variables and the outcome in the same year, we cannot establish the direction of these relationships, and a country’s HIV incidence may itself shape the service coverage and financing patterns we treat as explanatory. Sixth, because the data are national aggregates, the associations we report describe countries and not individuals, and they may not hold for the people who use HIV services.

Future research should focus on three strategic areas. First, future research must incorporate novel data sources (such as Demographic and Health Surveys or Integrated Biological and Behavioural Surveillance Surveys microdata) to construct time-variant national proxies for PrEP uptake and the U=U metric (viral suppression rates) to test their direct association with the rate of new HIV infections over time. Second, longer panel studies should explore the OOP expenditure relationship over extended time lags to determine if financial burdens eventually translate into delayed transmission increases, particularly by constructing proxies for the financial strain on the most vulnerable populations. Third, alternative modelling techniques, such as the use of proxy variables or the Anderson-Hsiao estimator, could be investigated to potentially recover the effect of time-invariant variables like baseline health infrastructure while maintaining the Fixed-Effects methodology.

## Appendices

### Appendix 1: List of Relevant Variables from the GHO

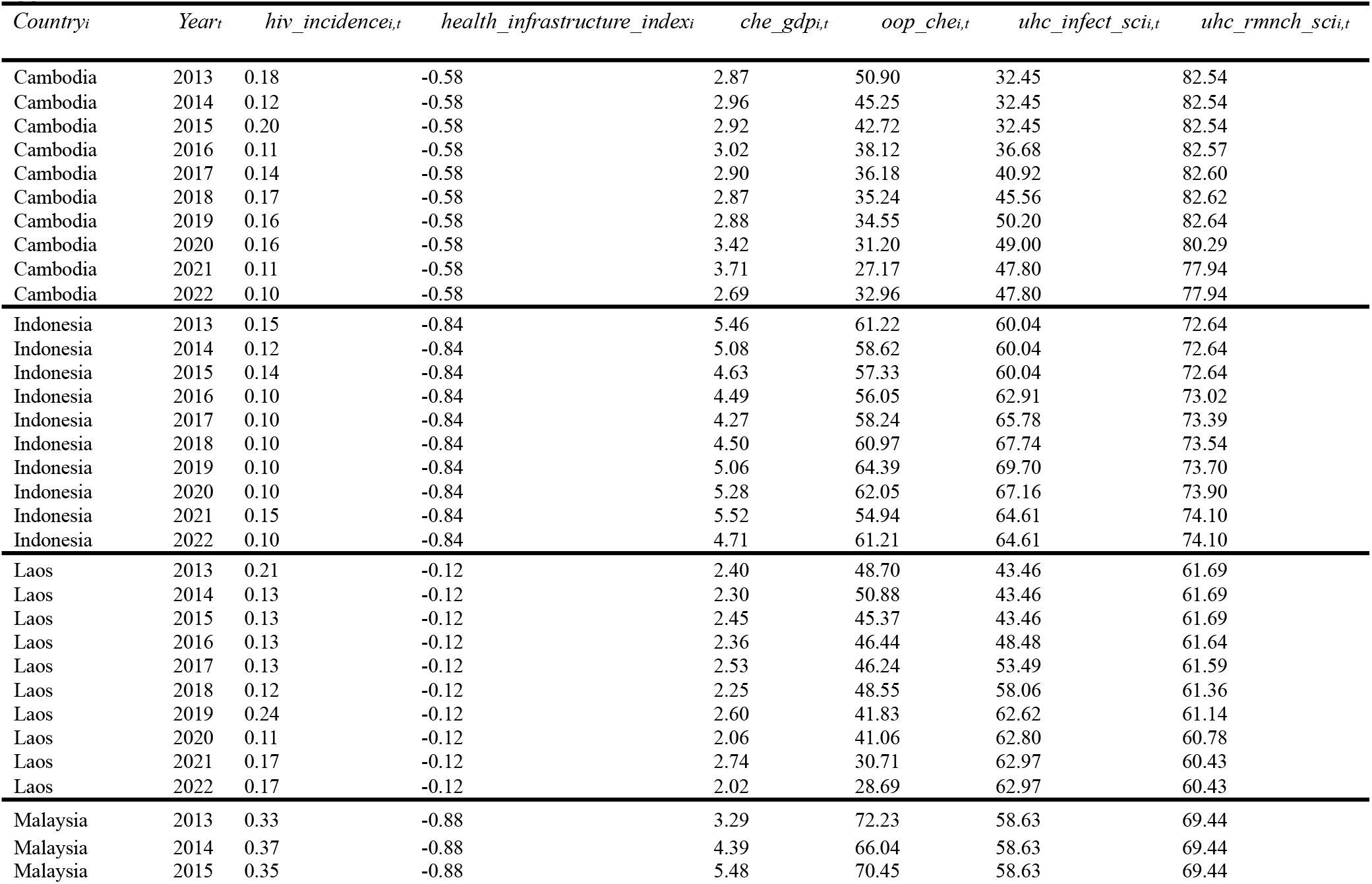
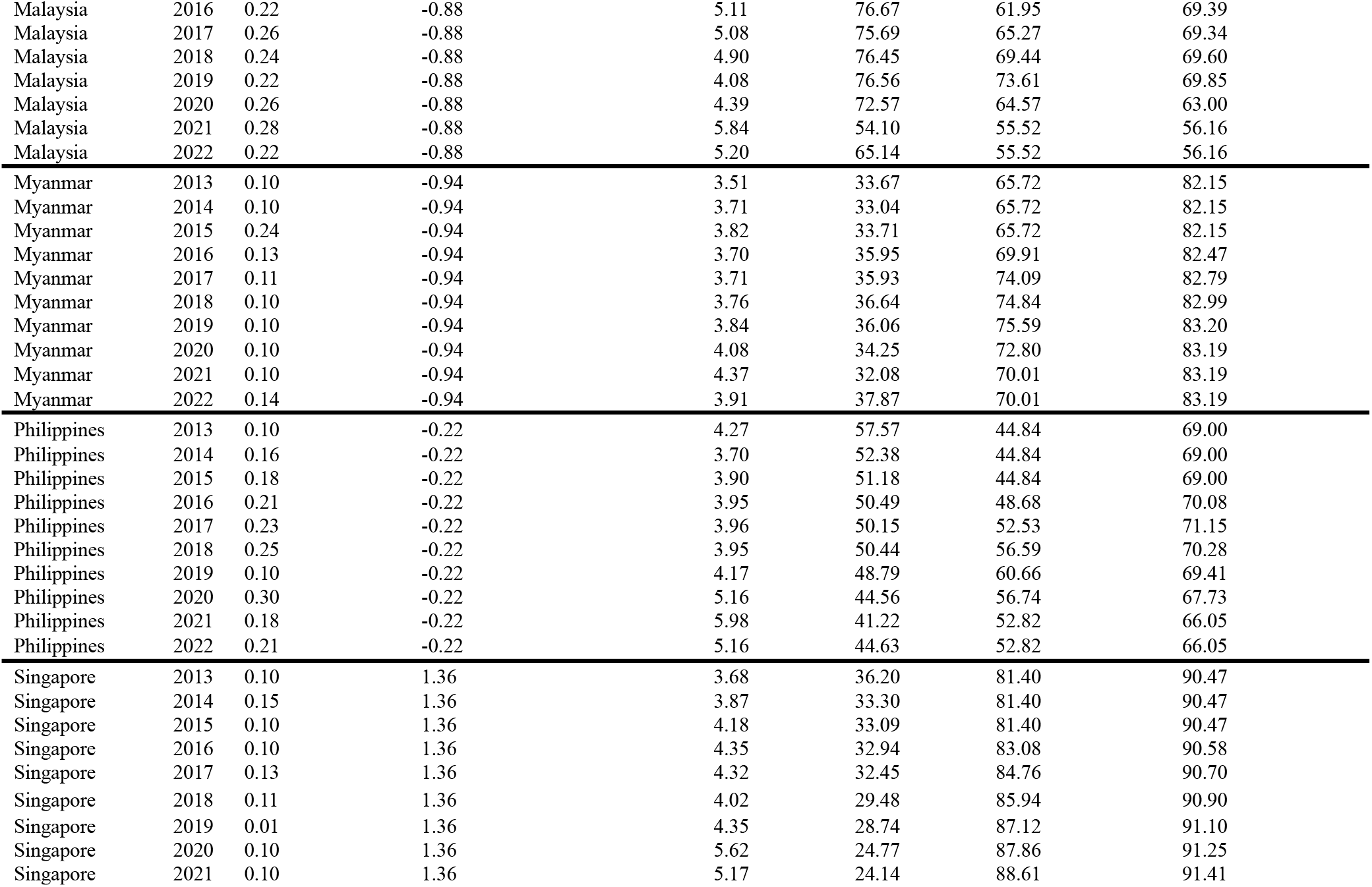
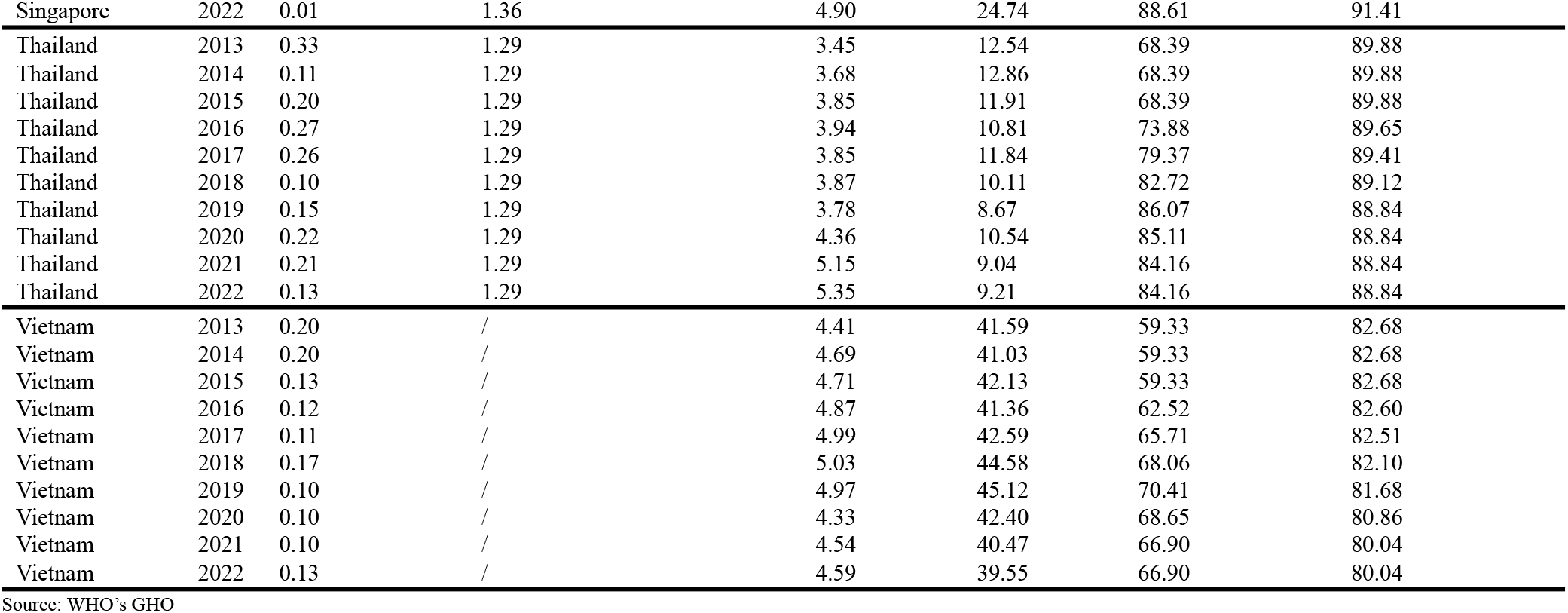

| *Country<sub>i</sub>* | *Year<sub>t</sub>* | *hiv_incidence<sub>i,t</sub>* | *health_infrastructure_index<sub>i</sub>* | *che_gdp<sub>i,t</sub>* | *oop_che<sub>i,t</sub>* | *uhc_infect_sci<sub>i,t</sub>* | *uhc_rmnh_sci<sub>i,t</sub>* |
| --- | --- | --- | --- | --- | --- | --- | --- |
| Cambodia | 2013 | 0.18 | -0.58 | 2.87 | 50.90 | 32.45 | 82.54 |
| Cambodia | 2014 | 0.12 | -0.58 | 2.96 | 45.25 | 32.45 | 82.54 |
| Cambodia | 2015 | 0.20 | -0.58 | 2.92 | 42.72 | 32.45 | 82.54 |
| Cambodia | 2016 | 0.11 | -0.58 | 3.02 | 38.12 | 36.68 | 82.57 |
| Cambodia | 2017 | 0.14 | -0.58 | 2.90 | 36.18 | 40.92 | 82.60 |
| Cambodia | 2018 | 0.17 | -0.58 | 2.87 | 35.24 | 45.56 | 82.62 |
| Cambodia | 2019 | 0.16 | -0.58 | 2.88 | 34.55 | 50.20 | 82.64 |
| Cambodia | 2020 | 0.16 | -0.58 | 3.42 | 31.20 | 49.00 | 80.29 |
| Cambodia | 2021 | 0.11 | -0.58 | 3.71 | 27.17 | 47.80 | 77.94 |
| Cambodia | 2022 | 0.10 | -0.58 | 2.69 | 32.96 | 47.80 | 77.94 |
| Indonesia | 2013 | 0.15 | -0.84 | 5.46 | 61.22 | 60.04 | 72.64 |
| Indonesia | 2014 | 0.12 | -0.84 | 5.08 | 58.62 | 60.04 | 72.64 |
| Indonesia | 2015 | 0.14 | -0.84 | 4.63 | 57.33 | 60.04 | 72.64 |
| Indonesia | 2016 | 0.10 | -0.84 | 4.49 | 56.05 | 62.91 | 73.02 |
| Indonesia | 2017 | 0.10 | -0.84 | 4.27 | 58.24 | 65.78 | 73.39 |
| Indonesia | 2018 | 0.10 | -0.84 | 4.50 | 60.97 | 67.74 | 73.54 |
| Indonesia | 2019 | 0.10 | -0.84 | 5.06 | 64.39 | 69.70 | 73.70 |
| Indonesia | 2020 | 0.10 | -0.84 | 5.28 | 62.05 | 67.16 | 73.90 |
| Indonesia | 2021 | 0.15 | -0.84 | 5.52 | 54.94 | 64.61 | 74.10 |
| Indonesia | 2022 | 0.10 | -0.84 | 4.71 | 61.21 | 64.61 | 74.10 |
| Laos | 2013 | 0.21 | -0.12 | 2.40 | 48.70 | 43.46 | 61.69 |
| Laos | 2014 | 0.13 | -0.12 | 2.30 | 50.88 | 43.46 | 61.69 |
| Laos | 2015 | 0.13 | -0.12 | 2.45 | 45.37 | 43.46 | 61.69 |
| Laos | 2016 | 0.13 | -0.12 | 2.36 | 46.44 | 48.48 | 61.64 |
| Laos | 2017 | 0.13 | -0.12 | 2.53 | 46.24 | 53.49 | 61.59 |
| Laos | 2018 | 0.12 | -0.12 | 2.25 | 48.55 | 58.06 | 61.36 |
| Laos | 2019 | 0.24 | -0.12 | 2.60 | 41.83 | 62.62 | 61.14 |
| Laos | 2020 | 0.11 | -0.12 | 2.06 | 41.06 | 62.80 | 60.78 |
| Laos | 2021 | 0.17 | -0.12 | 2.74 | 30.71 | 62.97 | 60.43 |
| Laos | 2022 | 0.17 | -0.12 | 2.02 | 28.69 | 62.97 | 60.43 |
| Malaysia | 2013 | 0.33 | -0.88 | 3.29 | 72.23 | 58.63 | 69.44 |
| Malaysia | 2014 | 0.37 | -0.88 | 4.39 | 66.04 | 58.63 | 69.44 |
| Malaysia | 2015 | 0.35 | -0.88 | 5.48 | 70.45 | 58.63 | 69.44 |
| Malaysia | 2016 | 0.22 | -0.88 | 5.11 | 76.67 | 61.95 | 69.39 |
| Malaysia | 2017 | 0.26 | -0.88 | 5.08 | 75.69 | 65.27 | 69.34 |
| Malaysia | 2018 | 0.24 | -0.88 | 4.90 | 76.45 | 69.44 | 69.60 |
| Malaysia | 2019 | 0.22 | -0.88 | 4.08 | 76.56 | 73.61 | 69.85 |
| Malaysia | 2020 | 0.26 | -0.88 | 4.39 | 72.57 | 64.57 | 63.00 |
| Malaysia | 2021 | 0.28 | -0.88 | 5.84 | 54.10 | 55.52 | 56.16 |
| Malaysia | 2022 | 0.22 | -0.88 | 5.20 | 65.14 | 55.52 | 56.16 |
| Myanmar | 2013 | 0.10 | -0.94 | 3.51 | 33.67 | 65.72 | 82.15 |
| Myanmar | 2014 | 0.10 | -0.94 | 3.71 | 33.04 | 65.72 | 82.15 |
| Myanmar | 2015 | 0.24 | -0.94 | 3.82 | 33.71 | 65.72 | 82.15 |
| Myanmar | 2016 | 0.13 | -0.94 | 3.70 | 35.95 | 69.91 | 82.47 |
| Myanmar | 2017 | 0.11 | -0.94 | 3.71 | 35.93 | 74.09 | 82.79 |
| Myanmar | 2018 | 0.10 | -0.94 | 3.76 | 36.64 | 74.84 | 82.99 |
| Myanmar | 2019 | 0.10 | -0.94 | 3.84 | 36.06 | 75.59 | 83.20 |
| Myanmar | 2020 | 0.10 | -0.94 | 4.08 | 34.25 | 72.80 | 83.19 |
| Myanmar | 2021 | 0.10 | -0.94 | 4.37 | 32.08 | 70.01 | 83.19 |
| Myanmar | 2022 | 0.14 | -0.94 | 3.91 | 37.87 | 70.01 | 83.19 |
| Philippines | 2013 | 0.10 | -0.22 | 4.27 | 57.57 | 44.84 | 69.00 |
| Philippines | 2014 | 0.16 | -0.22 | 3.70 | 52.38 | 44.84 | 69.00 |
| Philippines | 2015 | 0.18 | -0.22 | 3.90 | 51.18 | 44.84 | 69.00 |
| Philippines | 2016 | 0.21 | -0.22 | 3.95 | 50.49 | 48.68 | 70.08 |
| Philippines | 2017 | 0.23 | -0.22 | 3.96 | 50.15 | 52.53 | 71.15 |
| Philippines | 2018 | 0.25 | -0.22 | 3.95 | 50.44 | 56.59 | 70.28 |
| Philippines | 2019 | 0.10 | -0.22 | 4.17 | 48.79 | 60.66 | 69.41 |
| Philippines | 2020 | 0.30 | -0.22 | 5.16 | 44.56 | 56.74 | 67.73 |
| Philippines | 2021 | 0.18 | -0.22 | 5.98 | 41.22 | 52.82 | 66.05 |
| Philippines | 2022 | 0.21 | -0.22 | 5.16 | 44.63 | 52.82 | 66.05 |
| Singapore | 2013 | 0.10 | 1.36 | 3.68 | 36.20 | 81.40 | 90.47 |
| Singapore | 2014 | 0.15 | 1.36 | 3.87 | 33.30 | 81.40 | 90.47 |
| Singapore | 2015 | 0.10 | 1.36 | 4.18 | 33.09 | 81.40 | 90.47 |
| Singapore | 2016 | 0.10 | 1.36 | 4.35 | 32.94 | 83.08 | 90.58 |
| Singapore | 2017 | 0.13 | 1.36 | 4.32 | 32.45 | 84.76 | 90.70 |
| Singapore | 2018 | 0.11 | 1.36 | 4.02 | 29.48 | 85.94 | 90.90 |
| Singapore | 2019 | 0.01 | 1.36 | 4.35 | 28.74 | 87.12 | 91.10 |
| Singapore | 2020 | 0.10 | 1.36 | 5.62 | 24.77 | 87.86 | 91.25 |
| Singapore | 2021 | 0.10 | 1.36 | 5.17 | 24.14 | 88.61 | 91.41 |
| Singapore | 2022 | 0.01 | 1.36 | 4.90 | 24.74 | 88.61 | 91.41 |
| Thailand | 2013 | 0.33 | 1.29 | 3.45 | 12.54 | 68.39 | 89.88 |
| Thailand | 2014 | 0.11 | 1.29 | 3.68 | 12.86 | 68.39 | 89.88 |
| Thailand | 2015 | 0.20 | 1.29 | 3.85 | 11.91 | 68.39 | 89.88 |
| Thailand | 2016 | 0.27 | 1.29 | 3.94 | 10.81 | 73.88 | 89.65 |
| Thailand | 2017 | 0.26 | 1.29 | 3.85 | 11.84 | 79.37 | 89.41 |
| Thailand | 2018 | 0.10 | 1.29 | 3.87 | 10.11 | 82.72 | 89.12 |
| Thailand | 2019 | 0.15 | 1.29 | 3.78 | 8.67 | 86.07 | 88.84 |
| Thailand | 2020 | 0.22 | 1.29 | 4.36 | 10.54 | 85.11 | 88.84 |
| Thailand | 2021 | 0.21 | 1.29 | 5.15 | 9.04 | 84.16 | 88.84 |
| Thailand | 2022 | 0.13 | 1.29 | 5.35 | 9.21 | 84.16 | 88.84 |
| Vietnam | 2013 | 0.20 | / | 4.41 | 41.59 | 59.33 | 82.68 |
| Vietnam | 2014 | 0.20 | / | 4.69 | 41.03 | 59.33 | 82.68 |
| Vietnam | 2015 | 0.13 | / | 4.71 | 42.13 | 59.33 | 82.68 |
| Vietnam | 2016 | 0.12 | / | 4.87 | 41.36 | 62.52 | 82.60 |
| Vietnam | 2017 | 0.11 | / | 4.99 | 42.59 | 65.71 | 82.51 |
| Vietnam | 2018 | 0.17 | / | 5.03 | 44.58 | 68.06 | 82.10 |
| Vietnam | 2019 | 0.10 | / | 4.97 | 45.12 | 70.41 | 81.68 |
| Vietnam | 2020 | 0.10 | / | 4.33 | 42.40 | 68.65 | 80.86 |
| Vietnam | 2021 | 0.10 | / | 4.54 | 40.47 | 66.90 | 80.04 |
| Vietnam | 2022 | 0.13 | / | 4.59 | 39.55 | 66.90 | 80.04 |

Source: WHO's GHO

### Appendix 2: Variance Inflation Factor (VIF)

| Variable | VIF | 1/VIF |
| --- | --- | --- |
| *health_infrastructure_index<sub>i</sub>* | 3.08 | 0.32 |
| *che_gdp<sub>i,t</sub>* | 2.46 | 0.41 |
| *oop_che<sub>i,t</sub>* | 2.30 | 0.43 |
| *uhc_infect_sci<sub>i,t</sub>* | 2.14 | 0.47 |
| *uhc_rmnch_sci<sub>i,t</sub>* | 1.53 | 0.65 |
| Mean VIF | 2.30 |  |

## Supplementary Material

### (3) Fixed-Effects Model (Lagged)

The Fixed-Effects model removes all time-invariant country heterogeneity (α_i_). It tests how changes in the lagged explanatory variables within a country affect the current outcome.

*Fixed-Effects Model: hiv_incidence*_*i,t*_ *= β*_*0*_ *+ β*_*2*_*che_gdp*_*i,t-1*_ *+ β*_*3*_*oop_che*_*i,t-1*_ *+ β*_*4*_ *uhc_infect_sci*_*i,t-1*_ *+ β*_*5*_*uhc_rmnch_sci*_*i,t-1*_ *+ α*_*i*_ *+ ϵ*_*it*_

Assumptions for the Fixed-Effects estimation:

1. **Strict exogeneity (lagged)**: The idiosyncratic error term (ϵ_it_) is uncorrelated with the lagged regressors (X_1,t-1_) across all time periods.
2. **No perfect collinearity**: The lagged regressors (X_1,t-1_) must vary over time within each country.
3. **Time-invariant variable omission**: The time-invariant variable (Z_i_) is absorbed into α_i_ and is omitted from the estimation.
4. **Error structure (clustered)**: The use of *vce(cluster country_id)* ensures that the inference is valid, as it explicitly assumes:
  a. Errors are correlated within each country (autocorrelation is allowed).
  b. Errors may have different variances across countries (heteroscedasticity is allowed).
  c. Errors are independent between countries (clusters).

### (4) Random-Effects Model (Lagged)

The Random-Effects model estimates the effect using a weighted average of the within- and between-country variation. It allows for the estimation of the time-invariant variable (Z_i_).

*Random-Effects Model: hiv_incidence*_*i,t*_ *= β*_*0*_ *+ β*_*1*_*health_infrastructure_index*_*i*_ *+ β*_*2*_*che_gdp*_*i,t-1*_ *+ β*_*3*_*oop_che*_*i,t-1*_ *+ β*_*4*_ *uhc_infect_sci*_*i,t-1*_ *+ β*_*5*_*uhc_rmnch_sci*_*i,t-1*_ *+ (α*_*i*_ *+ ϵ*_*it*_)

Assumptions for the Random-Effects estimation:

1. **No correlation**: The country-specific error term (α_i_) must be uncorrelated with all lagged and time-invariant explanatory variables (X_1,t-1_ and Z_i_).
2. **Lag structure**: It assumes the relationship between the predictors and the outcome is fully captured by the one-period lag structure.
3. **Error structure (clustered)**: Similar to the Fixed-Effects model, *vce(cluster country_id)* provides robust inference by allowing for autocorrelation and heteroscedasticity within each country cluster.

**Supplementary Table 1:** Regression Results for HIV Incidence (Lagged Explanatory Variables & Clustered Errors)

| Variable | Fixed-Effects model |  | Random-Effects model |  |
| --- | --- | --- | --- | --- |
|  | coefficient | SE | coefficient | SE |
| *health_infrastructure_index<sub>i</sub>* | / |  | -0.0121 | 0.0272 |
| *che_gdp<sub>i,t-1</sub>* | -0.0255 | 0.0154 | -0.0206* | 0.0117 |
| *oop_che<sub>i,t-1</sub>* | -0.0020 | 0.0016 | -0.0001 | 0.0006 |
| *uhc_infect_sci<sub>i,t-1</sub>* | -0.0009 | 0.0008 | -0.0003 | 0.0008 |
| *uhc_rmch_sci<sub>i,t-1</sub>* | 0.0039 | 0.0022 | -0.0002 | 0.0008 |
| constant | 0.1043 | 0.1358 | 0.2767*** | 0.0972 |
| N (Observations) | 81 |  | 72 |  |
| N (Groups) | 9 |  | 8 |  |
| F-statistic/ Wald $\chi^2$ | F(4,8) = 1.97 | | $\chi^2(5) = 6.86$ | |
| p-value | 0.1921 |  | 0.2316 |  |
| Sargan-Hansen C-statistic | $\chi^2(4) = 17.87$ | $p < 0.001$ | | |

Note: \*\*\*, \*\*, \* denotes significance at the 0.1%, 1%, and 5% level, respectively.
Note: The lower N in the Random-Effects model reflects the exclusion of Vietnam, for which the *health\_infrastructure\_index* could not be estimated due to missing facility density data throughout the study period.

## Acknowledgements

N/A.

## Author Contributions

Conceptualisation: J.H.; Methodology: J.H.; Data curation and formal analysis: J.H.; Writing – original draft: J.H.; Writing – review and editing: J.H. and A.S.; Supervision: A.S.

## Ethics Statement

Ethics approval was not required for this study as all data were drawn from publicly available, anonymised, aggregated surveillance datasets published by the WHO’s GHO.

## Data Availability

All data used in this study are publicly available from the WHO’s GHO API at: https://www.who.int/data/gho/info/gho-odata-api.

